# Multimodal neuroimaging data from a 5-week heart rate variability biofeedback randomized clinical trial

**DOI:** 10.1101/2022.12.28.22283798

**Authors:** Hyun Joo Yoo, Kaoru Nashiro, Jungwon Min, Christine Cho, Noah Mercer, Shelby L. Bachman, Padideh Nasseri, Shubir Dutt, Shai Porat, Paul Choi, Yong Zhang, Vardui Grigoryan, Tiantian Feng, Julian F. Thayer, Paul Lehrer, Catie Chang, Jeffrey A. Stanley, Vasilis Z. Marmarelis, Shrikanth Narayanan, Daniel A. Nation, Jessica Wisnowski, Mara Mather

**Affiliations:** University of Southern California; University of Groningen; University of California, Los Angeles; University of California, Irvine; Rutgers University; Vanderbilt University; Wayne State University School of Medicine

## Abstract

We present data from the Heart Rate Variability and Emotion Regulation (HRV-ER) randomized clinical trial testing effects of HRV biofeedback. Younger (N = 121, ages 18-31, 61 female) and older (N = 72, ages 55-80, 45 female) participants completed baseline magnetic resonance imaging (MRI) including T_1_-weighted, functional MRI (fMRI), pulsed continuous arterial spin labeling (PCASL), and proton magnetic resonance spectroscopy (^1^H MRS). They also completed an emotion regulation task during fMRI. During fMRI scans, physiological measures (blood pressure, pulse, respiration, and end-tidal CO_2_) were continuously acquired. Participants were randomized to either increase heart rate oscillations or decrease heart rate oscillations during daily sessions. After 5 weeks of HRV biofeedback, they repeated the baseline measurements in addition to a few new measures (ultimatum game fMRI, training mimicking during blood oxygen level dependent (BOLD) and PCASL fMRI). Participants also wore a wristband sensor to estimate sleep time. Psychological assessment comprised three cognitive tests as well as ten questionnaires related to emotional well-being. Data is publicly available via the OpenNeuro data sharing platform.

## Background & Summary

Heart rate variability (HRV) is one of the most consistent correlates of psychological and emotional well-being and stress^1-3^. However, it is not just random variation in the interval between heartbeats that is associated with well-being. In healthy resting people, heart rate is tonically suppressed by signals transmitted via the vagus nerve. This suppression of heart rate is stronger when exhaling than when inhaling^4^, and it is “vagal HRV” or the high frequency oscillations in heart rate in response to breathing that are most strongly associated with positive well-being (or low negative affect or stress). Spending time every day breathing at a pace of around 10 seconds per breath (a pace that induces resonance with the baroreflex and so induces high oscillations in heart rate) while getting biofeedback on heart rate oscillatory activity can enhance well-being^5,6^. This suggests that heart rate oscillatory activity serves as more than a readout of the integrity of the brain’s autonomic regulatory systems. Short bouts of high heart rate oscillations may stimulate these regulatory systems, enhancing their function^7^. To test this hypothesis, in a randomized clinical trial (ClinicalTrials.gov NCT03458910), we scanned younger and older participants while at rest and while doing an emotion regulation task both before and after five weeks of daily practice sessions in which they received heart rate variability biofeedback to either increase or decrease heart rate oscillations.

Initial studies using heart rate variability biofeedback yielded promising results and there has been a significant growth in interest in this intervention^5,6^. Compared with most prior HRV-biofeedback studies, our study has a larger N and a more extensive set of outcome measures. It is also unique among HRV-biofeedback studies in having all of the following features: functional and structural brain outcome measures, a well-matched active comparison group, inclusion of two age groups, and heart rate data available from each practice session. Thus, these Heart Rate Variability and Emotion Regulation (HRV-ER) clinical trial data should be a rich source for a variety of secondary analyses, including those investigating individual-difference factors that affect responses to HRV-biofeedback, examination of age differences in response to the intervention, and specific patterns of brain changes in response to the intervention. Furthermore, the baseline pre-intervention data could be relevant for potential secondary analyses unrelated to heart rate biofeedback. For instance, the larger N than seen in most fMRI emotion regulation studies allows for individual difference comparisons, especially given all the additional physiological data collected from each participant. In addition, this study includes data not typically available in public datasets, such as PCASL, a turbo spin echo (TSE) structural sequence targeting the locus coeruleus, and biochemical measurements using proton magnetic resonance spectroscopy (^1^H MRS), allowing for unique secondary analyses not previously feasible.

## Methods

### Power Considerations for sample size

No prior studies had examined effects of these interventions on brain function so we were unable to estimate effect sizes based on prior neuroimaging data. We elected to power our study to detect medium or larger effect sizes. Our main planned statistical comparisons were repeated-measures ANOVAs with within-between interactions. For these, a total sample size of 46 would give 90% power to detect moderate effect sizes of f = .25 with *α* = .05, given an assumed correlation among the repeated measures of .5 (1). We also planned to examine within-subject change within each of the conditions. A sample size of 44 in each group would give 90% power to detect within-group change effect sizes of d = .5 in a two-tailed t-test with *α* = .05 (1). Thus, we aimed for an N = 100 completion rate across the two groups for each age group to be able to accommodate potential exclusions for movement during imaging or other data quality issues.

### Participants

We recruited 121 younger participants aged between 18 and 35 years and 72 older participants aged between 55 and 80 years via the USC Healthy Minds community subject pool, a USC online bulletin board, Facebook and flyers between February 2018 and March 2020 (see Fig. 1 for drop-out rates per condition; note that older adult enrollment was cut short by the COVID pandemic). Participants provided informed consent approved by the University of Southern California (USC) Institutional Review Board. Participants were recruited in waves of approximately 20 participants at a time, each of whom was assigned to a small group of 3-6 people. Groups met for weekly lab visits at the same time and day each week. After group assignments of a wave were complete, we assigned each group to one of two conditions involving daily biofeedback that aimed to increase heart rate oscillations (Osc+ condition) or decrease heart rate oscillations (Osc-condition). We used blocked randomization across small groups to maintain balanced numbers of each condition. We did this by determining how many groups were assigned to a condition; for example, if 2 out of 5 groups in a previous wave were assigned the Osc+ condition, then 3 out of 5 groups were assigned that condition in the next wave. Then we randomly allocated those conditions to groups. One research staff member who was blinded to participants and small group assignment generated the random numbers and assigned the conditions to each small group. The study utilized a single-blinded design; the consent document did not mention that there were two conditions and participants in both conditions were told that we were interested in how training to control heart rate might influence emotional health and the functions of brain regions involved in emotion regulation. Upon completing the study, participants were paid for their participation and received bonus payments based on their individual and group performances (incentives for training were the same across conditions; see Supplementary Information for more details). Prospective participants were screened and excluded for major medical, neurological, or psychiatric illnesses. We excluded people who had a disorder that would impede performing the HRV biofeedback procedures (e.g., coronary artery disease, angina, cardiac pacemaker), who currently were training using a relaxation, biofeedback or breathing practice, or were on any psychoactive drugs other than antidepressants or anti-anxiety medications. We included people who were taking antidepressant or anti-anxiety medication and/or attending psychotherapy only if the treatment had been ongoing and unchanged for at least three months and no changes were anticipated. Gender, education, age, and race did not differ significantly in the two conditions.

**Fig. 1.**
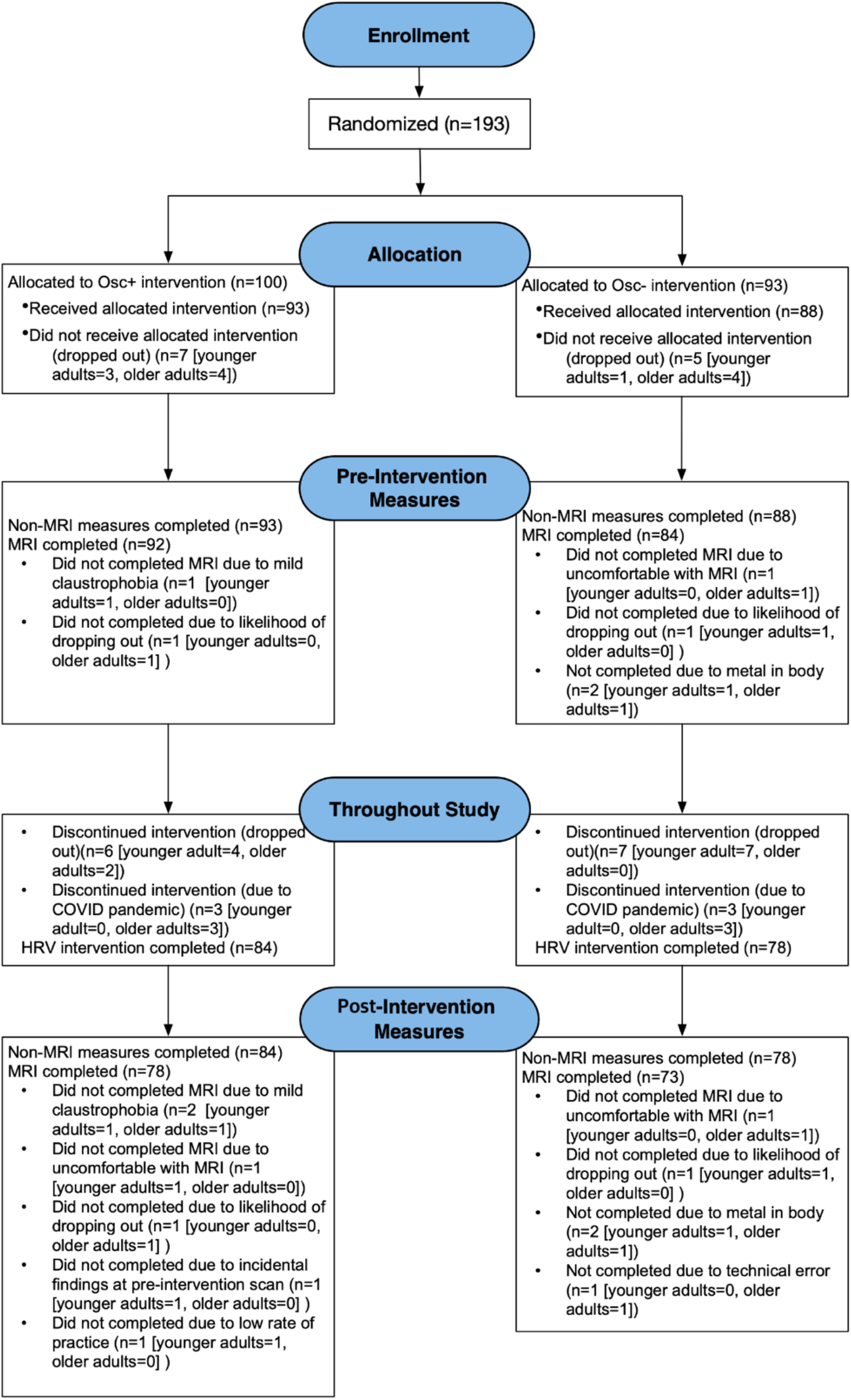
CONSORT flow diagram showing participant flow through each stage of the randomized controlled trial.

### Overview of 7-week protocol schedule

The study protocol involved seven weekly lab visits and five weeks of home biofeedback training (Fig. 2). Each lab visit began with questionnaires assessing mood and anxiety. The first lab visit involved non-MR baseline measurements, including several questionnaires. The second lab visit involved the baseline MR session, followed by the first biofeedback calibration and training session (see below for details). The weekly lab visits (except for weeks with MR sessions) were run in small groups in which participants shared their experiences and tips about biofeedback training with other participants from the same condition, while 1-2 researchers facilitated the discussion. Outside the lab, participants used a customized social app to communicate with other group members and researchers about their progress on daily biofeedback training. The Week 6 lab visit repeated the assessments from the first lab visit. The final (7th) lab visit repeated the baseline MR session scans in the same order. Additional training-session scans were collected at the end of the scan protocol. Finally, after the scan, participants completed a post-study questionnaire. Table 1 provides detailed information about the measurement at each time point.

**Fig. 2.**
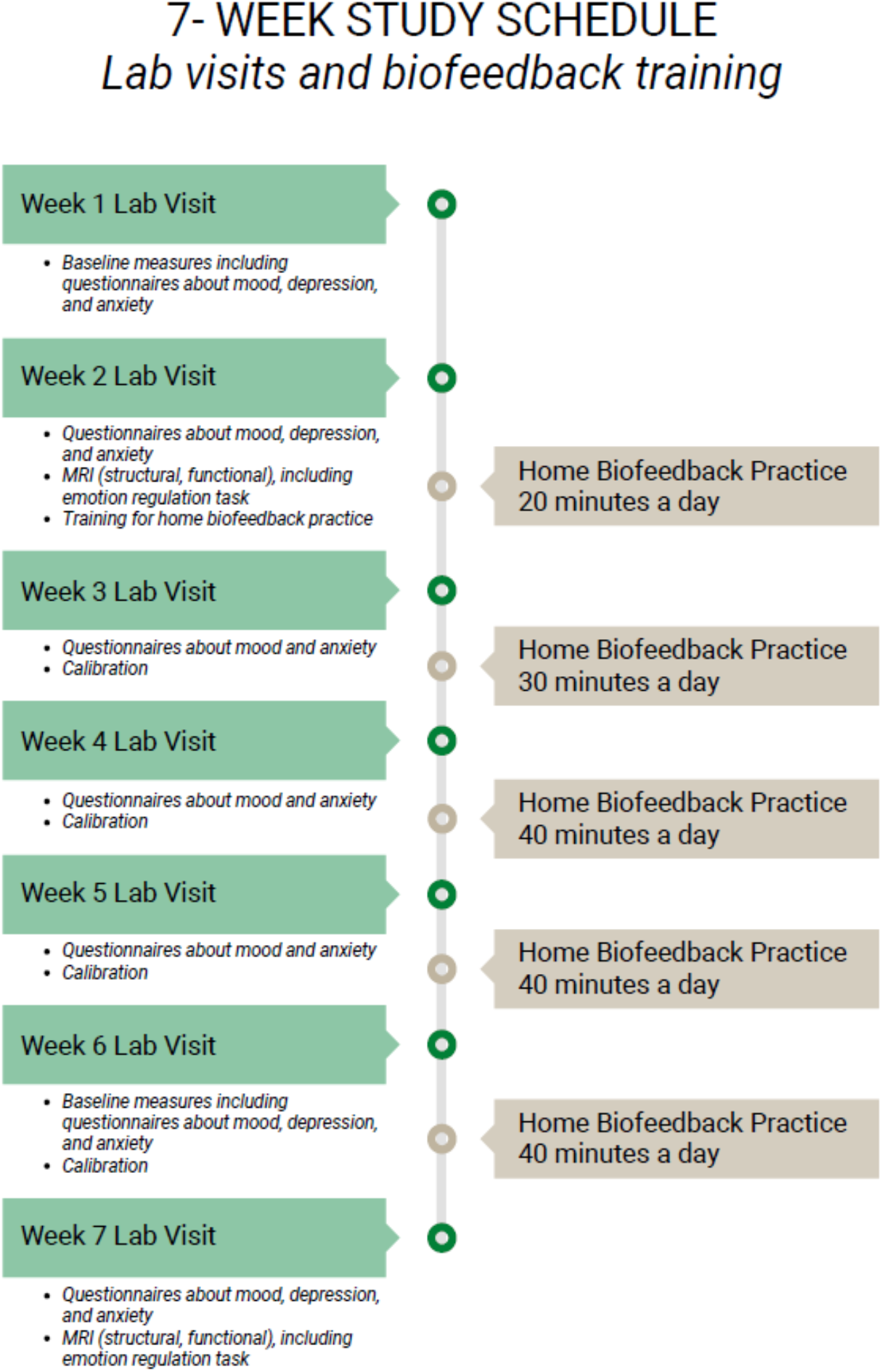
Study 7-week schedule.

**Table 1.** Measurement at each time point.

| Category | Data type | Week1 | Week2 | Week3 | Week4 | Week5 | Week6 | Week7 | Measurement |
| --- | --- | --- | --- | --- | --- | --- | --- | --- | --- |
| Demographics | Demographics | * |  |  |  |  |  |  | •Self-report |
| Questionnaire | Emotion-weekly (POMS, SAI) | * | * | * | * | * | * | * | •Self-report |
|  | Emotion-4 times (CESD, TAI) | * | * |  |  |  | * | * | •Self-report |
|  | Altruism |  | * |  |  |  |  |  | •Self-report |
|  | others-pre/post (FFMQ) | * |  |  |  |  | * |  | •Self-report |
| Cognitive task | NIH toolbox: Cognition | * |  |  |  |  | * |  | •score |
|  | SART | * |  |  |  |  | * |  | •score<br>•response time |
|  | Picture memory task |  |  |  | * | * |  |  | •encoding (week 4)<br>•recognition (week 5)<br>•recall (week 5) |
| Heart rate data | calibration |  | * | * | * | * | * | * | •Inter-beat-interval data from pulse |
|  | Home training |  | * | * | * | * | * |  | •inter-beat-interval data from pulse |
| Stress task | Physiological data | * |  |  |  |  | * |  | •ECG, respiration, GSR, continuous blood pressure |
|  | Behavioral data | * |  |  |  |  | * |  | •score |
| MR Imaging | Functional-resting |  | * |  |  |  |  | * | •brain imaging data<br>•physiological data |
|  | Functional-resting ASL |  | * |  |  |  |  | * | •brain imaging data<br>•physiological data |
|  | Functional-Emotion regulation |  | * |  |  |  |  | * | •brain imaging data<br>•physiological data<br>•event data |
|  | Anatomical-T<sub>1</sub> |  | * |  |  |  |  | * | •brain imaging data |
|  | <sup>1</sup>H MRS |  | * |  |  |  |  | * | •brain biochemistry data |
|  | Anatomical-TSE |  | * |  |  |  |  | * | •brain imaging data (locus coeruleus) |
|  | Functional-UG |  |  |  |  |  |  | * | •brain imaging data<br>•physiological data<br>•event data |
|  | Functional-training mimicking |  |  |  |  |  |  | * | •brain imaging data<br>•physiological data<br>•event data |
|  | Functional-training mimicking ASL |  |  |  |  |  |  | * | •brain imaging data<br>•physiological data<br>•event data |

#### Biofeedback training

##### Osc+ group

Participants wore an ear sensor to measure their pulse. They viewed real-time heart rate biofeedback while breathing in through the nose and out through the mouth in synchrony with a visual pacer. The software^8^ provided a summary ‘coherence’ score for participants calculated as peak power/(total power – peak power). Peak power was determined by finding the highest peak within the range of 0.04 – 0.26 Hz and calculating the integral of the window 0.015 Hz above and below this highest peak. Total power was computed for the 0.0033 – 0.4 Hz range.

During the second lab visit, we introduced participants to the device and had them complete five minutes of paced breathing at 9 s, 10 s, 11 s, 12 s, and 13 s per breath, which approximately corresponds with 6.5, 6, 5.5, 5 and 4.5 breaths/min in Lehrer et al.^9^ Next, we computed various aspects of the oscillatory dynamics for each breathing pace using Kubios HRV Premium 3.1 software^10^ and assessed which breathing pace had the most of the following characteristics: highest LF power, the highest maximum LF amplitude peak on the spectral graph, highest peak-to-trough amplitude, cleanest and highest-amplitude LF peak, highest coherence score and highest root mean squared successive differences (RMSSD). Participants were then instructed to train at home with the pacer set to the frequency that appeared to best approximate their resonance frequency^9^ and to maximize their coherence scores.

During the third visit, they were asked to complete three 5-min paced breathing segments: the best condition from the last week’s visit, half a breath per minute faster, and half a breath slower than the best condition. They were then instructed to train the following week at the pace that appeared most likely to be a resonance frequency based on the characteristics listed above. In subsequent weekly visits, during 5-min training segments, they were asked to try out abdominal breathing and inhaling through the nose/exhaling through pursed lips as well as other strategies of their choice.

##### Osc- group

This condition was designed to be as similar as possible to the Osc+ condition, but with the opposing goal (to reduce heart rate oscillations). The same biofeedback ear sensor device was used in this condition and participants were asked to practice for the same amount of time. However, we created custom software to display a different set of feedback to the Osc- participants^11^. During each Osc- training session, a ‘calmness’ score was provided as feedback to the participants instead of the coherence score. The calmness score was calculated by multiplying the coherence score that would have been displayed in the Osc+ condition by -1 and adding 10 (an ‘anti-coherence’ score). The net result was that participants got more positive feedback (higher calmness scores) when their heart rate oscillatory activity in the 0.04 – 0.26 Hz range was low. The software also included a minor point penalty when heart rate was the lowest it had been in the most recent 15 s. Specifically, every 5 s, a local maximum IBI was set based on the maximum IBI from the past 15 s. If, at that point, the participant’s current IBI was longer than this local maximum, the calmness score displayed for the next 5 s was the anti-coherence score - 2. Naturally, most of the time, current IBI was lower than the local maximum, and in those cases, the calmness score was the anti-coherence score +1. Thus, there was a penalty in their calmness score for moments when their heart rate was slower than it had been in any of the past 15 s.

During the initial calibration session at the end of the second lab visit, each participant was introduced to the device and feedback and was asked to come up with five strategies to lower heart rate and heart rate oscillations. The participant was asked to wear the ear sensor and view real-time heart rate biofeedback while they tried each strategy for five minutes. We analyzed the data in Kubios and identified the best strategy as the one with the most of the following characteristics: lowest LF power, the minimum LF-amplitude peak on the spectral graph, lowest peak trough amplitude, multiple and lowest-amplitude LF-peak, highest calmness score and lowest RMSSD. Participants were then instructed to use this strategy to try to maximize their calmness scores in their home training sessions.

On the third visit, they were asked to select three strategies and try each out in a 5-min session. The strategy identified as best (based on the same characteristics used in the initial calibration session) was selected as the one to focus on during home training. In subsequent weekly visits, during 5-min training segments, they were again asked to try out strategies of their choice.

#### Heart rate data from lab calibration and home training

During lab calibration, pulse was measured using HeartMath emWave pro software with an infrared pulse plethysmograph (PPG) ear sensor while participants sat in a chair with knees at a 90 degrees angle and both feet flat on the floor. The pulse wave was recorded with a sampling rate of 370 Hz. Interbeat interval (IBI) data was extracted after eliminating ectopic beats and other artifactual signals through a built-in process in emWave pro. On each home training session, pulse was measured using the same devices and software used for calibration sessions. An IBI data file was saved on a study-provided laptop and transferred to the lab server after completing each session.

### Questionnaires

#### Emotion questionnaires

During each lab visit, participants completed the profile of mood states (POMS)^12^ and the State Anxiety Inventory (SAI)^13^. We used the 40-item version of POMS. Participants reported how much each item reflected how they felt at the moment using a scale from 1 (not at all) to 5 (extremely). Total mood disturbance was calculated by subtracting positive-item totals from negative-item totals. A constant value (i.e. 100) was added to the total mood disturbance to eliminate negative scores. The SAI measures state anxiety using 20 statements. Participants indicated how they felt at the moment on a scale from 1 (not at all) to 4 (very much so). In Weeks 1, 2, 6 and 7, we also administered the Trait Anxiety Inventory (TAI)^13^ and the Center for Epidemiological Studies Depression Scale (CES-D) in Weeks 1, 2, 6 and 7^14^. The TAI measures trait anxiety using 20 statements, which participants rated on a 4-point scale from 1 (not at all) to 4 (very much so). The CES-D consists of 20 statements, which participants rated on a 4-point scale from 0 (rarely) to 3 (most or all of the time). At Week 1 and Week 6 lab visits, participants completed six additional emotion questionnaires. We assessed trait mindfulness using the 20-item version of the Five Facet Mindfulness Questionnaire (FFMQ)^15^. Participants rated each item using a scale from 1 (never) to 5 (very often/always true). We also administered the Smith Relaxation States Inventory 3 (SRSI3)^16^ to assess various aspects of stress, relaxation, meditation, and mindfulness. Participants completed state and disposition versions of the SRSI3 each consisting of 38 items. The state version asks how you “feel right now” on a 6-point scale from 1 (not at all) to 6 (maximum). The disposition version asks how often you experience relaxation states and stress states. We slightly modified the disposition version and asked how often each item has been experienced “in the past month” on a 6-point scale from 1 (rarely or never, less than once a month) to 6 (a lot, more than once a day). We calculated SRSI3 scores based on the Relaxation/Meditation/Mindfulness (RMM) Tracker/SRSI3 Manual v9.9.2020, which includes 34 items for scoring^17^. We measured the extent and severity of fatigue using the 11-item Chalder Fatigue Scale (CFQ 11)^18^ on a 4-point scale from 0 (less than usual) to 3 (much more than usual) or 0 (better than usual) to 3 (much worse than usual). We also administered the 10-item Emotion Regulation Questionnaire (ERQ)^19^, which is designed to measure tendency to regulate emotions in two ways (cognitive reappraisal and expressive suppression) on a 7-point scale from 1 (strongly disagree) to 7 (strongly agree). Participants also completed a self-efficacy version of the 10-item ERQ, which asks how “capable” they are of regulating their emotions on the same 7-point scale. In addition, perceived stress was measured using the NIH Toolbox Perceived Stress Survey^20^, a 10-item version of the Perceived Stress Scale^21^. Participants rated the frequency of stressful experiences and the extent to which they felt strained or overloaded during the past month (e.g., How often have you felt nervous and “stressed”? How often have you felt difficulties were piling up so high that you could not overcome them?) on a five-point scale, ranging from Never (1) to Very Often (5); higher scores correspond to greater perceived stress. Score on the perceived stress scale has been calculated as the mean score. All the rest are calculated as sum scores.

#### Altruism questionnaire

Participants completed the Altruism scale questionnaire^22^ during their lab visits on the second week of intervention. Altruism scale questionnaire is a self-report scale with 20 items each describing an altruistic behavior (e.g., “I have done volunteer work for a charity” and “I have delayed an elevator and held the door open for a stranger). Participants were instructed to rate the frequency of engaging in these behaviors on a 5-point scale (1 = never; 2 = once, 3 = more than once; 4 = often; 5 = very often). Higher scores in this scale correspond with higher altruistic tendencies.

#### Demographics and post-study questionnaire

The week-1 visit, participants completed questionnaires including basic demographics, clinical history including medications.

After the Week-7 post-intervention scan, participants completed a questionnaire surveying their experience during the study. They provided self-ratings of difficulty of daily heart rate biofeedback training, level of effort to complete the training, expectations of the training impact on well-being, and likelihood of continuing the training after the study’s conclusion.

### Stress task

During the week 1 and week 6 lab visits, participants completed a task designed to assess reactivity to and recovery from acute stress. The task consisted of several phases: a 4-minute baseline resting phase, a stress phase, and a 4-minute recovery resting phase. Younger participants completed two computerized tasks during the stress phase: a Paced Auditory Serial Addition Task (PASAT) and a modified Stroop color-word matching task. Seven older participants completed a pilot version of the stress phase which included both the PASAT and Stroop tasks. But the rest of older participants completed only the Stroop task. Descriptions of the PASAT and Stroop tasks are provided below. To enhance the socially evaluative nature of the tasks, participants were told that their performance would be evaluated by the experimenter and compared with that of other participants. In addition, auditory feedback was provided during each task, with a buzzer sound played for incorrect or missed responses and a bell sound played for correct responses^23^. During the baseline and recovery resting phases, participants sat with their feet resting flat on the ground, their hands resting in a prone position on a flat table, and their eyes open.

Physiological signals were recorded during all phases of the stress task using a BIOPAC MP160 system at a sampling frequency of 2 KHz. Electrocardiogram (ECG) and respiration signals were sent to the MP160 with a BioNomadix wireless transmitter. ECG signals were measured using a Lead II configuration and disposable, pre-gelled Ag/AgCl electrodes (EL501). Respiration signals were assessed with the BIOPAC Respiratory Effort Transducer, which involved a belt being placed around the lower rib cage to measure changes in chest circumference. Blood pressure signals were measured from the non-dominant arm with a BIOPAC noninvasive blood pressure monitoring system (NIBP100D). For electrodermal activity recordings, disposable, pre-gelled Ag/AgCl electrodes (EL507) were attached to the palmar side of the medial phalange of the fourth and fifth fingers of each participant’s non-dominant hand (as the index and middle fingers were used for continuous blood pressure recordings); leads were connected to a BIOPAC GSR100C module. Raw physiological signals were split into segments corresponding to the phases of the stress task.

#### Paced Auditory Serial Addition Task (PASAT)

During the PASAT, participants were shown a series of digits. They were instructed to add each digit to the digit shown previously and enter the sum using the keyboard^24^. The sum always ranged between 1-20. The PASAT consisted of 30 trials, and participants completed 4 practice trials prior to beginning the task.

#### Modified Stroop Task

On each trial of the modified Stroop color-word matching task, participants first saw one of two instructions: “color” or “meaning,” followed by a color word (RED, GREEN, or BLUE) presented in one of three colors incongruent with its meaning (either red, green or blue)^25^. Participants were instructed to press a key corresponding to either the color or meaning of the word stimulus, depending on the instruction shown immediately before the word. The task consisted of 20 trials, and participants completed 4 practice trials before beginning the task.

### Cognitive tasks

#### The National Institutes of Health Toolbox Cognitive Battery

The National Institutes of Health (NIH) Toolbox Cognitive Battery is a component of the NIH Toolbox for Assessment of Neurological and Behavioral Function (www.nihtoolbox.org;^26,27^) that comprises extensively validated computer-administered cognitive tests for use across childhood and adolescence, early adulthood, and old age. We administered the NIH-Toolbox cognitive battery using an iPad app on a 9.7 inch iPad Air 2. The Flanker Test, the List Sorting Working Memory (LSWM) Test, and the Pattern Comparison Processing Speed (PCPS) Test were administered to evaluate attention and executive function, working memory, and processing speed, respectively.

In addition to raw scores and/or reaction time, the NIH-toolbox cognitive battery generates computed scores and three types of standard scores for each subtest: uncorrected standard scores, age-corrected standard scores, and fully adjusted scores that account for age, education, gender, and race/ethnicity.

##### The Flanker inhibitory control and attention test

The Flanker test is a version of the Eriksen flanker task derived from the Attention Network Test^28^. On each trial, a central directional target (arrows for ages 8 and older) is flanked by similar stimuli on the left and right. The participant chooses the direction of the central stimulus. On 12 congruent trials, the flankers face the same direction as the target. On 8 incongruent trials, they face the opposite direction. A scoring algorithm integrates accuracy and reaction time, a measure more relevant to adult performance on this task, yielding computed scores from 0 to 10. There are 20 trials for ages 8 and older, and the test duration is about 4 min. Score is based on an algorithm derived from both accuracy and reaction time if the former is >80%. If less than 80%, score is accuracy. The test takes approximately 3 minutes to administer.

##### The List Sorting Working Memory Test

This task is an adaptation of Mungas’ List Sorting task from the Spanish and English Neuropsychological Assessment Scales^29,30^. In this task, a list of stimuli is presented both visually (picture) and auditorily (recording of a one-word description of the stimulus) on a computer monitor, one at a time at a rate of 2 sec per stimulus, and participants are required to repeat all the stimuli back to the examiner in order of increasing real-world size, from smallest to largest. In the first phase of the test (i.e., the 1-List phase), participants are first shown a list with two items drawn from a single category (i.e., food). If participants are correct on this 2-item list, the number of items in the list presented on the next trial increases by one item, up to a total of seven items per list (i.e., list length ranges from a 2-item list to a 7-item list, for a total of six levels of list length). If participants err on a trial at a given list length, they receive another trial with the same number of items in the list; if they err on that trial, this phase of the test is discontinued. Following the 1-list phase, all participants proceed to the second phase of the test (the 2-list phase), in which they see lists of items drawn from two different categories (i.e., food and animals). Participants are instructed to reorder and repeat the stimuli first from one category, then the other, in order of size within each category. Lists in the 2-list phase start with a 2-item list and increase in number of items in the same way as in the 1-list phase. For both phases, for each list length, participants receive a score of 2 points if they are correct on the first trial. A second trial at a given list length is only administered when participants fail the first trial. Participants receive a score of 1 point only for a given list length if they fail the first trial at that list length but pass the second trial. Test scores consist of combined total trials correct on the 1-list and 2-list phases of the task. The test takes approximately 7 minutes to administer.

##### The Pattern Comparison Processing Speed Test

The Pattern Comparison Processing Speed Test is modeled after Salthouse’s Pattern Comparison Task^31^. This test requires participants to identify whether two visual patterns are the “same” or “not the same” (responses were made by pressing a “yes” or “no” button). Patterns are either identical or vary on one of three dimensions: color, adding/taking something away, or one versus many. Scores reflect the number of correct items (of a possible 130) completed in 90 s. The test takes approximately 3 minutes to administer.

#### Sustained Attention to Response Task (SART)

The SART^32^ was administered during Week 1 and Week 6 lab visits. During the task, participants were presented with a random series of single-digit numbers, ranging from 1 to 9. Participants were instructed to press the spacebar as soon as they saw each number other than 3. The task consisted of 225 trials where a single digit was presented for 250 ms with a 900-ms-lasting mask image between trials. The task took about 6 minutes and was based on the web-based Inquisit SART task developed by Millisecond Software.

#### Picture memory tasks

The emotional memory task was administered at the Week 4 and Week 5 lab visits. Seventy-two stimuli were selected from The Nencki Affective Picture System (NAPS)^33^, a database of realistic photographs that aim to induce positive, negative, or neutral emotional states. Stimuli were first counterbalanced by valence (24 each of positive, negative, and neutral); then two sets of 36 stimuli were created and counterbalanced by valence in each set (12 each of positive, negative, and neutral). Participants completed the task on the Qualtrics Survey platform.

At the Week 4 visit, one of the two sets of 36 stimuli were shown to participants by random assignment. After viewing each image, participants rated on a 1-9 scale how positive, negative, or neutral they found the images (1 = very negative to 9 = very positive, 5 = neutral). No time limit was imposed for participants to provide their ratings, and each image was shown on the screen for 4 seconds. Once all images in the set were rated, participants completed a free recall task, where they were asked to describe as many images they saw with as much detail as possible. There was no time limit imposed for the free recall.

At the Week 5 visit, participants first completed the same free recall task as in Week 4. Participants then viewed all 72 stimuli and for each image selected one of 3 response options: Remember, Know, and New. “Remember” was described as having a vivid memory of an image (i.e., it evoked thoughts or feelings when it was seen, recollections of something else that happened at that same moment, or where in sequence the image was). “Know” was described as being confident that the image was seen but that nothing specific related to thoughts, feelings, or experience was associated with the image. “New” was described as being confident that the image was not seen before. Each image was shown on the screen for 3 seconds, and there was no time limit for providing responses.

### Sleep time

Sleep and HRV derived from slow-wave sleep were measured with WHOOP wristbands^34^. Participants were provided WHOOP wristbands on the first day until the final week of the study. All participants were instructed to wear the wristband as close to 24 hours per day as possible. Sleep data metrics include, but are not limited to daily information of: hours in bed, hours of sleep, hours awake, hours of REM sleep, hours of deep sleep, hours of light sleep, number of disturbances, number of sleep cycles, hours of naps. Average heart rate per minute, resting heart rate, and heart rate variability were provided by WHOOP each day as well. WHOOP calculates sleep-derived HRV during the final 5 minutes of recorded deep sleep. Relative to gold standard polysomnography, WHOOP algorithms have been validated by independent researchers as having a 95% sensitivity for sleep, 68% sensitivity for deep sleep and 70% for REM sleep^35^.

### MRI/MRS data acquisition

#### MRI scan session order

In both the pre- and post-intervention MR sessions, scans were conducted in the following order: 1) resting-state during BOLD fMRI; 2) resting-state during pCASL; 3) emotion regulation task during fMRI; 4) T_1_-weighted structural scan; 5) magnetic resonance spectroscopy (MRS); and 6) T_1_-weighted TSE scan. The post-intervention session included three additional scans, which were performed between the ^1^H MRS and TSE scans in the following order: 1) ultimatum game task; 2) training-mimicking session during BOLD fMRI; and 3) training-mimicking session during pCASL. During both training-mimicking scans, participants engaged in their daily training without biofeedback (see below for details).

#### MRI scan parameters

We employed a 3T Siemens MAGNETOM Trio scanner with a 32-channel head coil at the USC Dana and David Dornsife Neuroimaging Center. T_1_-weighted 3D structural MRI brain scans were acquired pre and post intervention using a magnetization prepared rapid acquisition gradient echo (MPRAGE) sequence with TR = 2300 ms, TE = 2.26 ms, slice thickness = 1.0 mm, flip angle = 9°, field of view = 256 mm, and voxel size = 1.0 × 1.0 × 1.0 mm^3^, with 175 volumes collected (4:44 min). Functional MRI scans during resting-state, emotion-regulation, training and ultimatum game tasks were acquired using multi-echo echo-planar imaging sequence with TR= 2400 ms, TE 18/35/53 ms, slice thickness = 3.0 mm, flip angle = 75°, field of view = 240 mm, voxel size = 3.0 × 3.0 × 3.0 mm. We acquired 175 volumes (7:00 min) for the resting-state scan and training scan, 250 volumes (10:00 min) for the emotion-regulation task and 244 volumes (9:45 min) for the ultimatum game task. PCASL scans were acquired with TR = 3880 ms, TE = 36.48 ms, slice thickness = 3.0 mm, flip angle = 120°, field of view = 240 mm and voxel size = 2.5 × 2.5 × 3.0 mm^3^, with 12 volumes collected (3:14 min; 1st volume was an M0 image, 2nd volume was a dummy image that was discarded, and the remaining 10 volumes were five tag-control pairs) both during resting-state (pre and post intervention) and training-mimicking (post intervention) scans. This ASL approach provides high precision and signal-to-noise properties and has better test-retest reliability than pulsed or continuous ASL techniques^36^. The two-dimensional, multi-slice TSE scan was acquired with TR = 750 ms; TE = 12 ms; flip angle = 120°; bandwidth = 287 Hz/pixel; voxel size = 0.43 × 0.43 × 2.5 mm^3^, gap between slices = 1.0mm, 11 axial slices). The ^1^H MRS data were acquired using a single-voxel point-resolved spectroscopy (PRESS) sequence with an echo time of 35 ms and repetition times of 2.0 s from a 4.1 cm^3^ (1.6 × 1.6 × 1.6 cm^3^) voxel localized to the anterior portion of the anterior cingulate cortex. Axial, sagittal, and coronal orientations were assessed for accurate voxel placement. Metabolite spectra were acquired with water suppression (water saturation pulse with bandwidth of 50 Hz) and 128 signal averages. Additionally, 6 water reference scans were acquired. Total acquisition time, including prescans, was approximately 5 min, and all raw data were archived for processing offline.

#### Pre- and post-intervention BOLD resting-state scan

Participants were instructed to rest, breathe normally and look at the central white cross on the screen.

#### Pre- and post-intervention pCASL resting-state scan

To assess whether the intervention affected blood flow during rest, in both MR sessions participants completed a second short resting-state scan. Participants were instructed to rest while breathing normally with their eyes open. To make visual inputs like those viewed during the training scan (for our analyses comparing rest vs. training scans), we presented red and blue circles alternately at a random rate (see Training sessions during BOLD and pCASL section below). Participants were asked not to pay attention to these stimuli.

#### Training mimicking session during BOLD and pCASL

In the post-intervention scan session after the resting-state and emotion-regulation scans, participants completed their daily training without biofeedback during BOLD and pCASL scans. By this point, participants were well-trained, having each completed on average 57 training sessions at home. For the Osc+ group, a red and blue circle alternated at their resonance frequency. For example, if their resonance frequency was 12 sec, the red circle was presented for 6 sec followed by the blue circle for 6 sec. Participants were asked to breathe in with the red circle and breathe out with the blue circle. For the Osc- group, the stimuli were the same as for the Osc+ group; however, the red and blue circles alternated at a random rate and participants were told not to pay attention to them. See Fig. 3 for a visual representation of training mimicking session.

**Fig. 3.**
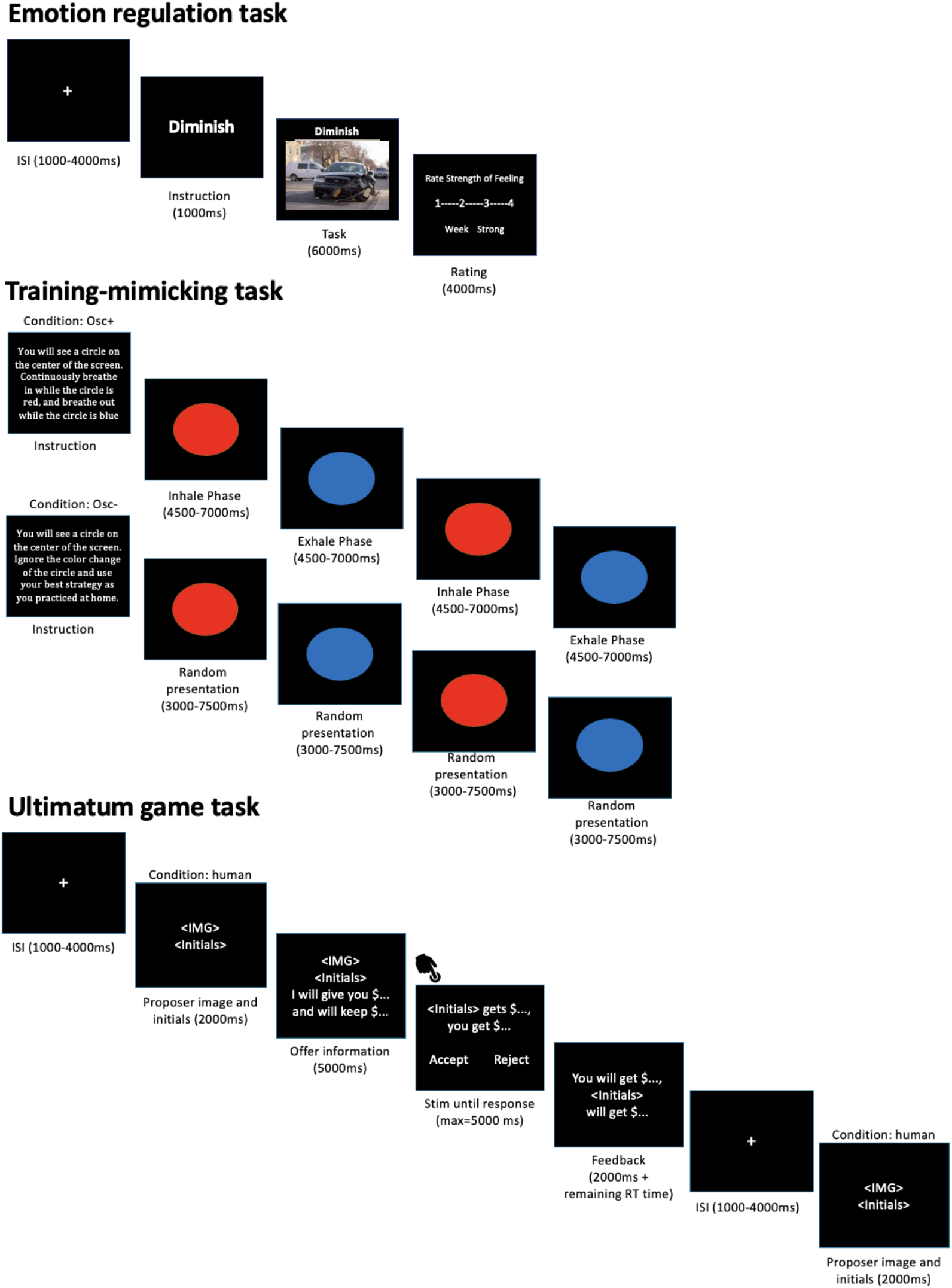
A visual representation of all experimental paradigms during task-based fMRI. ISI: inter-stimulus interval.

#### Emotion regulation task

Participants completed an emotion regulation task^37^ in the MRI scanner, which lasted for about 10 min. Each trial consisted of three parts: instruction (1s), regulation (6s), and rating (4s). First, participants were given one of three instructions: “view”, “intensify,” or “diminish.” Then, during the regulation phase, they saw a positive, neutral, or negative image. Finally, they were asked to rate the strength of the feeling they were experiencing on a scale ranging from 1 (very weak) to 4 (very strong). See Fig. 3 for a visual representation of emotion regulation task.

#### Ultimatum Game task

During the week 7 MRI scanning session, participants completed an Ultimatum Game (UG) task^38^. Before scanning, participants were instructed that in this task they would be presented by offers proposed by other participants of the study (with each player making 4 offers) or offers randomly generated by the computer. Participants had the options to accept or reject the offers. If they accepted an offer, the money would be split as proposed by the other player. If they rejected an offer, both players would receive nothing on that trial. Participants were told that at the end of the study, one of the trials in the game would be randomly selected and both them and the proposer for that trial would be paid based on the participant’s response. In order to enhance realism, participants also played the role of proposer and were asked to make offers to future participants. The task lasted for about 10 minutes and consisted of 36 trials. Out of these, 18 included fair and 18 included unfair offers. Fair offers ranged from 0.40 to 0.55 of the endowment whereas unfair offers were defined as ones ranging from 0.05 to 0.20 of the endowment. Each trial lasted 14 seconds. At the beginning of each trial, the face and initials of the proposer for that trial were presented for a duration of 2s. Then the offer proposed along with the face and initials of the proposer were shown for 5s. The decision period was then followed, during which participants had 5s to respond to the offer. Participants pressed one of two buttons on a button box to express their decision. Finally, the results screen was presented for a jittered duration of 2-6s. In between trials, a fixation cross was shown on screen which lasted between 1-4s. See Fig. 3 for a visual representation of Ultimatum Game task.

Younger and older adults completed two slightly different versions of the task. In younger adults’ version of the task, half of the human proposers were proposers that players knew, while the other half were strangers. For familiar proposers, pictures of participants’ group mates were used. In older adults’ version of the task, all human proposers were strangers.

#### Additional physiological measures during MRI

The physiological data collected during MRI scans include respiration, exhaled carbon dioxide (CO2), electrodermal activity, blood pressure, and heart rate. All the physiological data were collected at 10kHz sampling rate using Biopac MP150 Data Acquisition System with MR-compatible sensors and recorded with AcqKnowledge software 5.0. Respiration was measured using the breathing belt, TSD201 transducer and transferred to the Biopac RSP100C module to be 0.05-1Hz bandpass-filtered, amplified with 10 times of gain. Exhaled carbon dioxide (CO2) levels were measured using Philips NM3 Monitor (Model 7900) with nasal cannula and fed to Biopac MP150. The heart rate data were collected with a Nonin Medical 8600FO Pulse Oximeter and sent to Biopac MP150. The electrodermal activity was recorded using the Biopac GSR100C module. Blood pressure was measured using CareTaker device and software and recorded with Biopac MP150.

## Data Records

The following data are available on the OpenNeuro data sharing platform (https://openneuro.org/datasets/ds003823). The files were organized in Brain Imaging Data Structure (BIDS) format^39^ (version1.5.0; http://bids.neuroimaging.io). The BIDS provides a convention of fMRI data naming and organization in order to facilitate the transfer, storage, and sharing of neuroimaging data. The BIDS validation tool provided by OpenNeuro was used to ensure that the dataset followed the BIDS system.

At the root level of the dataset, participant demographic information, including sex, and handedness, and age group is provided in the participants.tsv file and these variables are further described in the accompanying data dictionary, participants.json. The participants.tsv file also indicates which of the different tasks, physiological data, and MRI scans are available for each participant at each time point. This information is organized into 33 columns containing “1” (data exist) or “0” (missing data) for all measures at each session (i.e., ses-pre_task-emotionRegulation; ses-post_task-emotionRegulation). Also, we organized the information about data quality in 15 columns containing “1” (recommend excluding) or “0” (recommend including) for MRI or physiological measures at each session.

We organized the rest of the participants’ data in three ways: phenotype, subject folders, and derivatives folder.

1. “Phenotype”: This folder includes files that list all participants’ scores on standardized tests at each time point and participants’ responses to emotion questionnaires at each time-point (with one row per participant)
2. “Sub-< ID > “: This folder contains participants’ MRI scan data, physiological measures, and behavioral measures. Inside the folder of each participant with data available (i.e., n = 193 for total participants and n = 162 for longitudinal data, see also Fig. 1 for detailed information), there are two subfolders, named “ses-pre”, “ses-post” that contain data collected during pre- and post-intervention sessions, respectively. Another two subfolders, named “ses-calibration”, “ses-home,” contain heart rate measures collected during in-lab calibration sessions and during home practice sessions, respectively. The last subfolder, named “beh” has individual data for the picture memory tasks, which were administered in Weeks 4 and 5. Inside “ses-pre” and “ses-post” folders, there are four subfolders named “anat”, “func”, “perf”, and “beh”. “Anat” folder contains T_1_-weighted structural images, “func” folder contains multi-echo BOLD scan data and the physiological data collected during those scans, “perf” folder contains pCASL scan data and the physiological data collected during those scans, and “beh” folder contains behavioral and physiological measures collected outside the scanner. Inside the “func” subfolder, there are files containing participant’s performance on the task (i.e., ‘events’ file), and physiological data for each task (i.e., ‘physio’ file) in addition to brain image files. The events file includes one row per trial, including onset time of each trial, duration of the event, trial type, response, response time, and the presented stimulus. Inside the “perf” subfolder, there are files containing (1) 10 tag & control acquisitions from the pCASL scan in a 4D file (“*_asl.nii”) and (2) an M0 calibration image from the pCASL scan in a 3D file (“*_m0scan.nii”). Fig. 4 provides an example of the BIDS data structure for one subject. Table 2 provides detailed information about the file name and the location for each measure at each time point.
3. “derivatives”: This folder contains MRS, mriqc, and freesurferQC folders. Inside the MRS folder, there is a “MRS_summary.tsv” file and individual subject folders. “MRS_summary.tsv” includes the individual metabolite concentration levels and quality metrics for all scans for all participants. Inside each subject folder, there are two subfolders, named “ses-pre”, “ses-post” including raw MRS data as IMA file format. The pre session included one MRS scan, which produced three ‘.IMA’ files. During the post session, some participants had two MRS scans; the first MRS scan occurred at the same point in the scan sequence as the pre MRS scan. The second, optional, MRS scan was completed after all other task scans were done. Inside the mriqc folder, there are multiple files for scan types; “group_T1w_mriqc.tsv” and “group_BOLD_mriqc_<task-name>.tsv” include the quality control metrics for the T_1_-weighted and functional (BOLD) MRI scans, respectively. Inside the freesurferQC folder, there is a freesurfer_QC.tsv file including Freesurfer quality metrics and outlier participants on these metrics flagged.

**Fig. 4.**
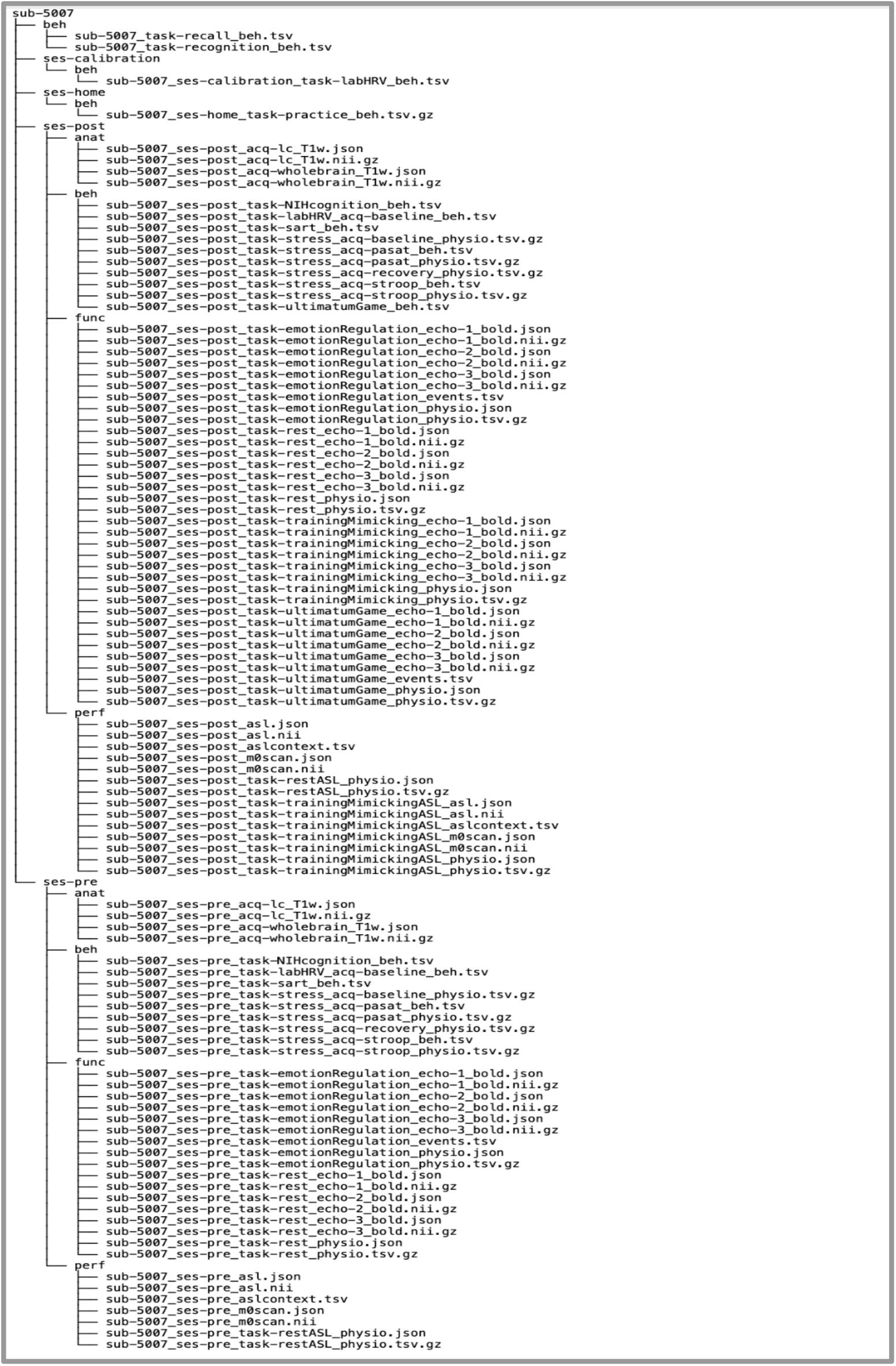
Example of the BIDS data structure for one participant. The data for subject 5007 are organized into five folders; two sessions for pre and post measurements, two sessions for HRV biofeedback data, one for calibration and the other for home training, and one last folder for behavioural data. While the data structure is consistent across subjects, there is some variation regarding data availability.

**Table 2.**
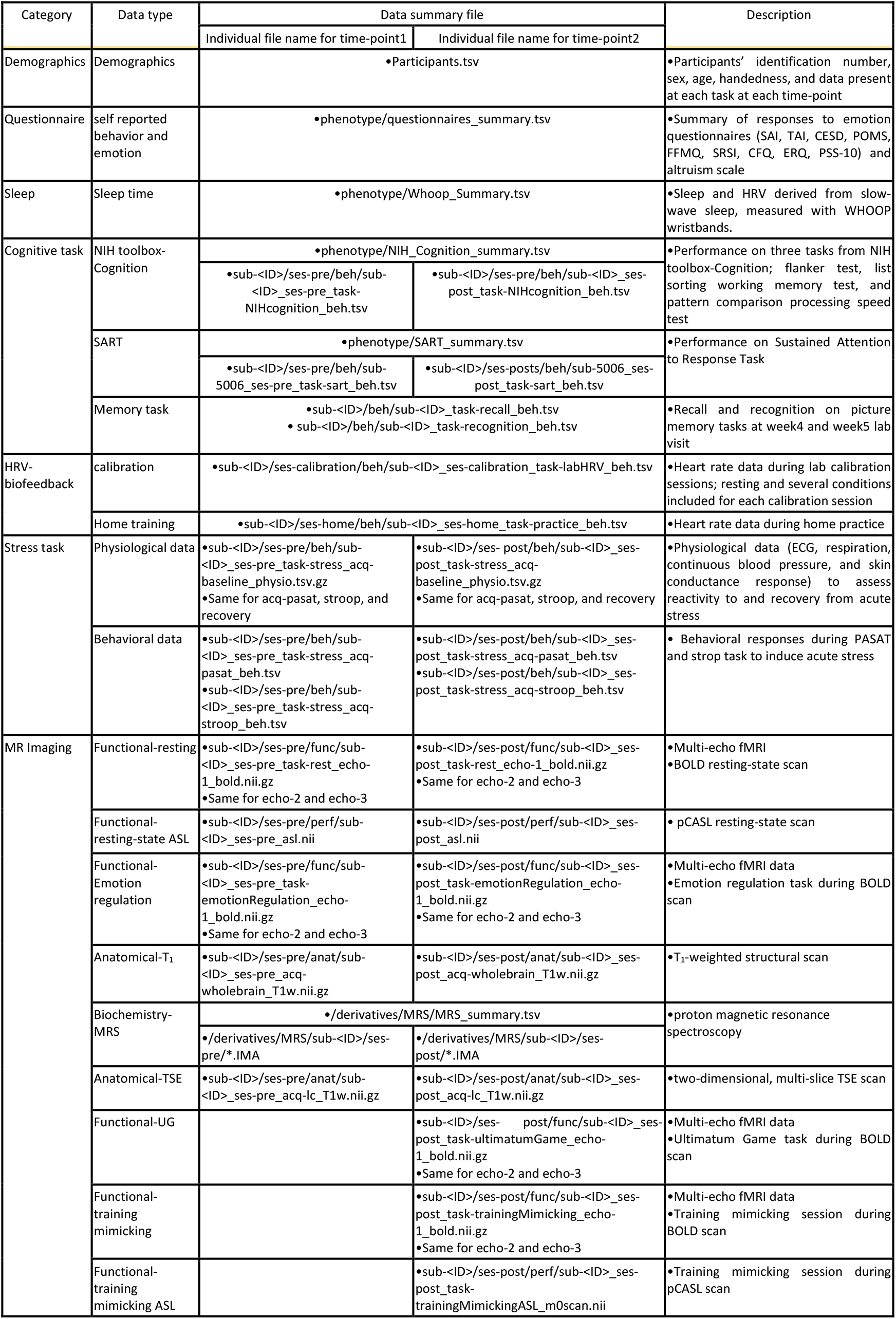
Summary of data file name and location.

## Technical Validation

In this section, we describe quality control metrics for each measure. We take a conservative approach for data exclusions. We generally did not exclude brain imaging data unless (1) the MRI scan session was interrupted by unexpected events (e.g., an earthquake or power outage), (2) an absence of a usable T_1_-weighted scan due to technical error or scan terminated by participants, or (3) incidental findings. Also, we did not exclude behavioral or physiological data unless (1) the task was interrupted by unexpected events (e.g., an earthquake or power outage) or (2) obvious sensor error or data input error due to a technical issue. But we applied quality control for data analyses and shared the results of quality control metrics in the derivative folder and quality control results in the participants.tsv file. This way, the future users of the datasets can use the quality-controlled data we recommend including, evaluate our quality control methods, or apply their own quality control methods on the datasets. Importantly, this places the responsibility for inclusion and exclusion of data in the hands of the users of the datasets.

### MRI Data quality assessment

The quality control metrics for the T_1_-weighted and functional (BOLD) MRI scans were computed by the MRIQC package, which outputs several quality control metrics as well as a report with visualizations of different aspects of the data. The quality control metrics for T_1_-weighted images are stored in the group_T1w.tsv in the derivatives/mriqc folder. The quality control metrics for the functional scans are stored in derivatives/mriqc folder.

#### T_1_-weighted scans

All T_1-_weighted scans were run through the MRIQC pipeline, which outputs several quality control metrics as well as a report with visualizations of different aspects of the data. All individual subject reports were visually checked for artifacts including reconstruction errors, failure of defacing, and segmentation. Defacing was considered to be successful if the 3D render did not contain more than one partial facial feature (eyes, nose, or mouth) and no brain tissue had been removed during defacing^40^. In Fig. 5, we visualize several quality control metrics related to the T_1_-weighted scans over two time-points (pre and post). MRIQC includes the signal-to-nose ratio (SNR) calculation proposed by Dietrich et al. ^41^, using the air background as a noise reference. Additionally, for images that have undergone some noise reduction processing, or the more complex noise realizations of current parallel acquisitions, a simplified calculation using the within tissue variance is also provided. Higher values indicate better quality. The contrast-to-noise ratio (CNR)^42^ is an extension of the SNR calculation to evaluate how separated the tissue distributions of GM and WM are. Higher values indicate better quality. The coefficient of joint variation (CJV) of GM and WM was proposed as objective function^43^ for the optimization of intensity non-uniformity (INU) correction algorithms. Higher values are related to the presence of heavy head motion and large INU artifacts. The entropy-focus criterion (EFC)^44^ uses the Shannon entropy of voxel intensities as an indication of ghosting and blurring induced by head motion. Lower values are better. Median INU is an index of spatial inhomogeneity. It estimates the location and spread of the bias field extracted^45^. The smaller spreads located around 1.0 are better. The white matter to maximum intensity ratio (WM2MAX) is the median intensity within the WM mask over the 95% percentile of the full intensity distribution, that captures the existence of long tails due to hyper-intensity of the carotid vessels and fat. Values should be around the interval [0.6, 0.8]^46^. In general, data quality appears consistent across time. All quality control metrics related to the T_1_-weighted scans for each participant, including those visualized in Fig. 5, are stored in the group_T1w_mriqc.tsv file in the derivatives/mriqc folder. Here we do not exclude any subjects based on IQMs, but subsequent researchers can use the available IQMs to exclude scans as they see fit.

**Fig. 5.**
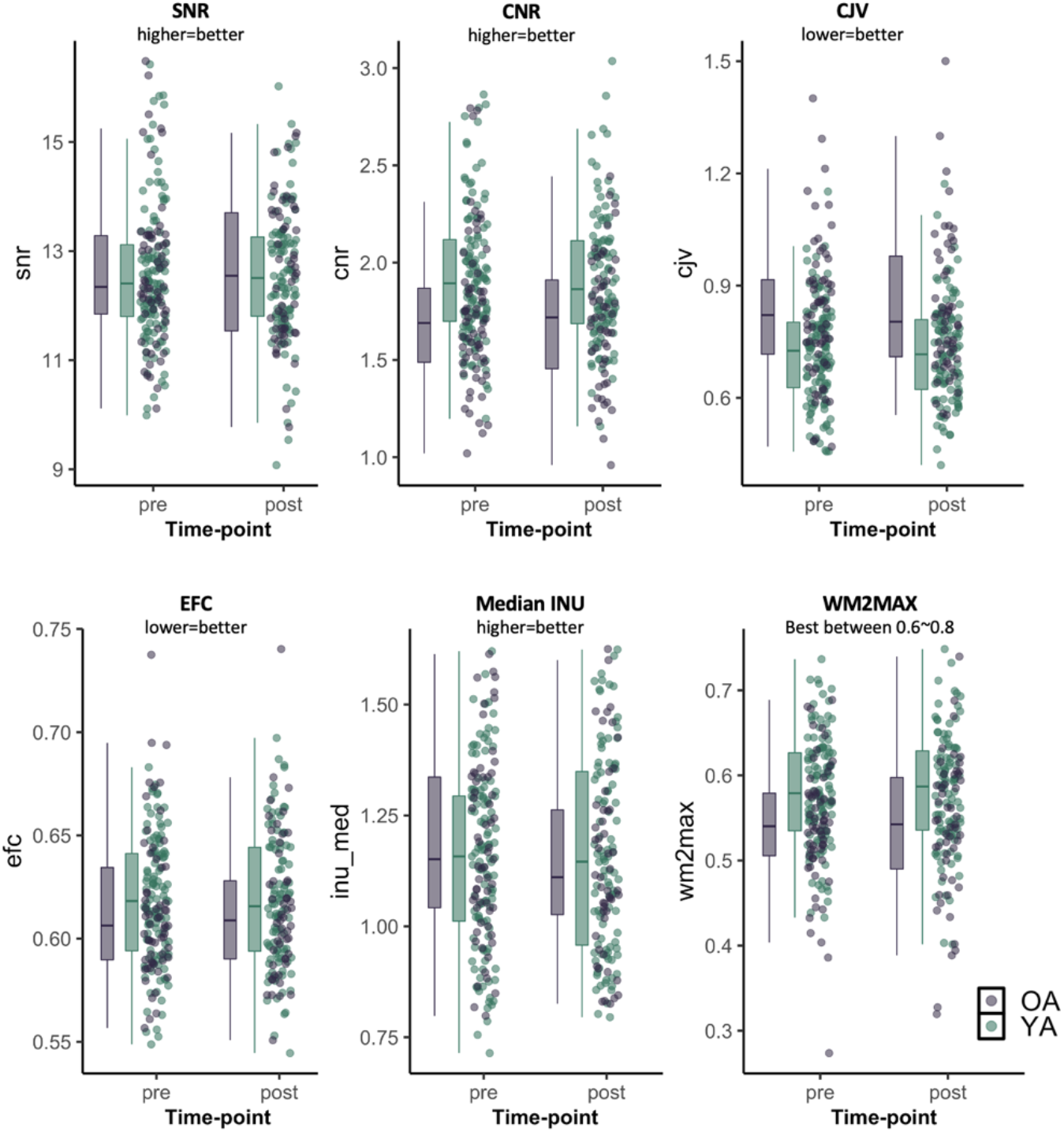
Quality control metrics related to the T_1_-weighted scans at each time-point. Quality control metrics related to the T_1_-weighted scans. SNR: signal-to-noise ratio; CNR: contrast-to-noise ratio; CJV: coefficient of joint variation, an index reflecting head motion and spatial inhomogeneity; EFC: entropy-focused criterion, an index reflecting head motion and ghosting; Median INU: intensity non-uniformity, an index of spatial inhomogeneity; WM2MAX: ratio of median white-matter intensity to the 95% percentile of all signal intensities

Each participant’s T_1_-weighted structural images were preprocessed using Freesurfer image analysis suite version 6.0 (http://surfer.nmr.mgh.harvard.edu/). Cortical reconstruction and volumetric segmentation were performed. Following initial preprocessing, we used the Freesurfer 6.0 image analysis suite longitudinal stream to automatically extract volume estimates^47^. After completing the longitudinal Freesurfer pipeline, we used automated measures computed by FreeSurfer of the contrast-to-noise ratio (the difference in signal intensity between regions of different tissue types and noise signal) and the Euler number (a metric of cortical surface reconstruction) to identify poor quality structural scans (Chalavi et al. 2012; Rosen et al. 2018). For analyses of volumetric change, we identified outliers (N = 4 for younger adults and N = 2 for older adults) who on a box-and-whisker plot were above Q3 + 3 × the interquartile range on either of these metrics on either pre or post images. Freesurfer quality metrics and the list of outliers are provided in the freesurfer_QC.tsv file in the derivatives/freesurferQC folder.

#### Functional (BOLD) scans

The functional (BOLD) scans were run through the MRIQC pipeline. The resulting reports were visually checked for artifacts including reconstruction errors, registration issues, and incorrect brain masks. In Fig. 6, we visualize several quality control metrics related to the functional scans across three echo times (e1 = 18ms, e2 = 35, and e3 = 53 ms) over two time-points (pre and post). Temporal SNR (tSNR) is a simplified interpretation of the tSNR definition^48^. The MRIQC pipeline provided the median value of the tSNR map calculated as, tSNR = ⟨S⟩_t_/σ_t_, where ⟨S⟩_t_ is the average BOLD signal (across time), and σ_t_ is the corresponding temporal standard-deviation map. Higher values are better when comparing scans at the same echo (differences across echo times are expected due to effects of echo time on BOLD contrast). Mean Framewise Displacement (FD) is a measure of subject head motion, which compares the motion between the current and previous volumes. Higher values indicate lower quality.

**Fig. 6.**
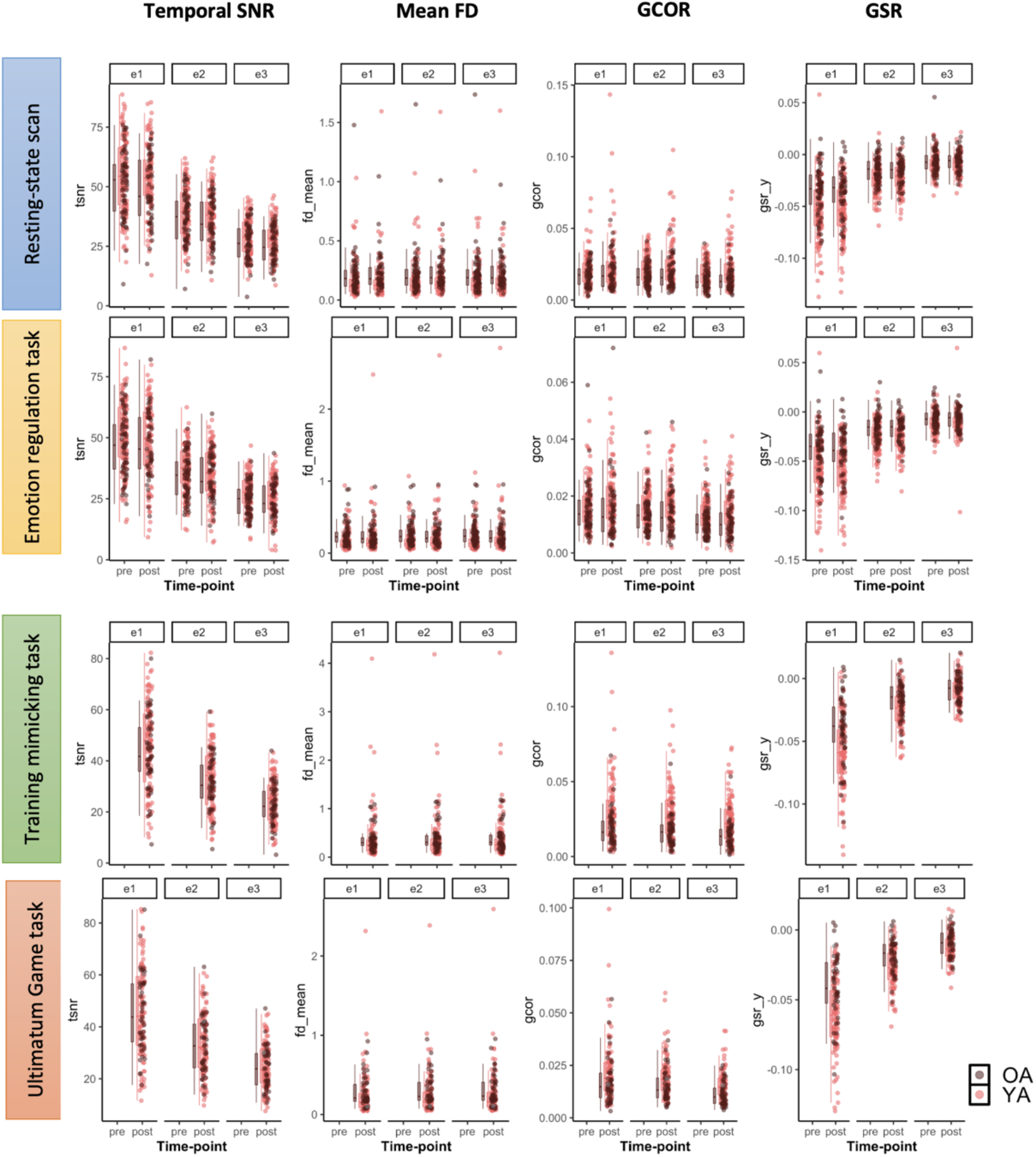
Quality control metrics related to the multi-echo functional (BOLD) scans at each time-point for resting-state scan, emotion regulation task scan, training mimicking task scan, and UG task. SNR: signal-to-noise ratio, an index of signal quality; FD: framewise displacement, an index of overall movement; GCOR: global correlation, an index of the presence of global signals; GSR: ghost-to-signal ratio, an index of ghosting along the phase-encoding axis.

Global Correlation (GCOR) is the average correlation of all pairs of voxel time series inside of the brain. GCOR measures differences between data due to motion/physiological noise/imaging artifacts as well as global neural fluctuations^49,50^. MRIQC measures ghost-to-signal ratio (GSR) along the x or y encoding axes. Higher values indicate lower quality. Like the T_1_-weighted quality control metrics, the functional quality metrics appear consistent across time. All quality control metrics related to the functional (BOLD) scans for each participant, including those visualized in Fig. 6, are provided in the group_BOLD_mriqc.tsv file in the derivatives/mriqc folder. Here we do not exclude any subjects based on IQMs, but subsequent researchers can use the available IQMs to exclude scans as they see fit.

#### MRS scans

Post-processing and quantification of the ^1^HMRS data were 100% automated^51^. For each ^1^H MRS spectra, the metabolites N-acetyl-aspartate (NAA), phosphocreatine plus creatine (PCr+Cr), trimethylamines [glycerophosphocholine plus phosphocholine (GP+CPC)], and myo-inositol, glutamate, and glutamine (as well as the less reliable metabolites, aspartate, gamma-aminobutyric acid, glutathione, lactate, n-acetylaspartylglutamate, scyllo-inositol and taurine) were quantified using the Linear Combination (LC) Model software^52^ with a simulated basis set for the a priori knowledge reflecting the acquisition parameters. An example of an individual MRS spectrum from the ^1^H MRS voxel placed in the anterior cingulate cortex is shown in Fig. 7. Freesurfer and FSL tools (FLIRT, FAST, MRI_VOLSYNTH, MRI_VOL2VOL) were used to tissue segment the T_1_-weighted images, which were then used to quantify the tissue fraction values within each voxel location.

**Fig. 7.**
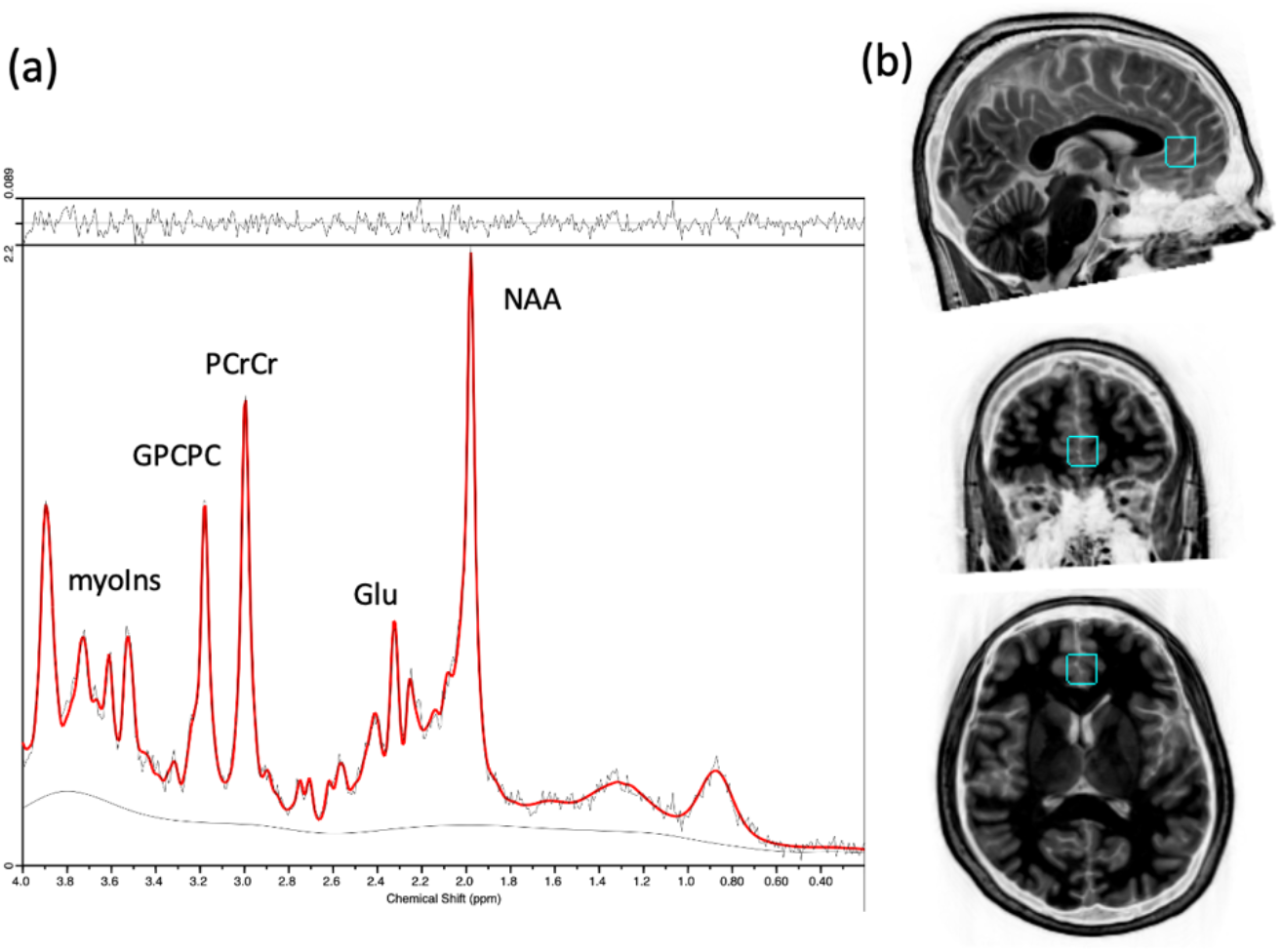
An example of an individual quantified MRS spectrum (a) and the sagittal, coronal and axial view of the MRS voxel placed in the anterior cingulate cortex, from top to bottom (b).

As quality control metrics for the MRS scans, we used SNR, line width reflecting the full width at half maximum (FWHM) of NAA, and Cramér-Rao lower bound (CRLB)^53^. Nine MRS spectra were rejected for poor quality (pre: 3, post-1st: 3, post-2nd: 3) due to extreme CRLB values and the distribution of quality metrics are visualized in Fig. 8 after removal of poor quality data. Also, all metabolite levels that have CRLB higher than 25% or another chosen threshold are tagged “outlier” with gray color in Fig. 8. But we included all data with quality tag in the shared data file, “MRS_summary.tsv”, for future users of the datasets to apply their own threshold on the datasets ^54^. The individual metabolite levels and quality metrics are provided for all scans for all participants in the MRS_summary.tsv file in the derivatives/MRS folder.

**Fig. 8.**
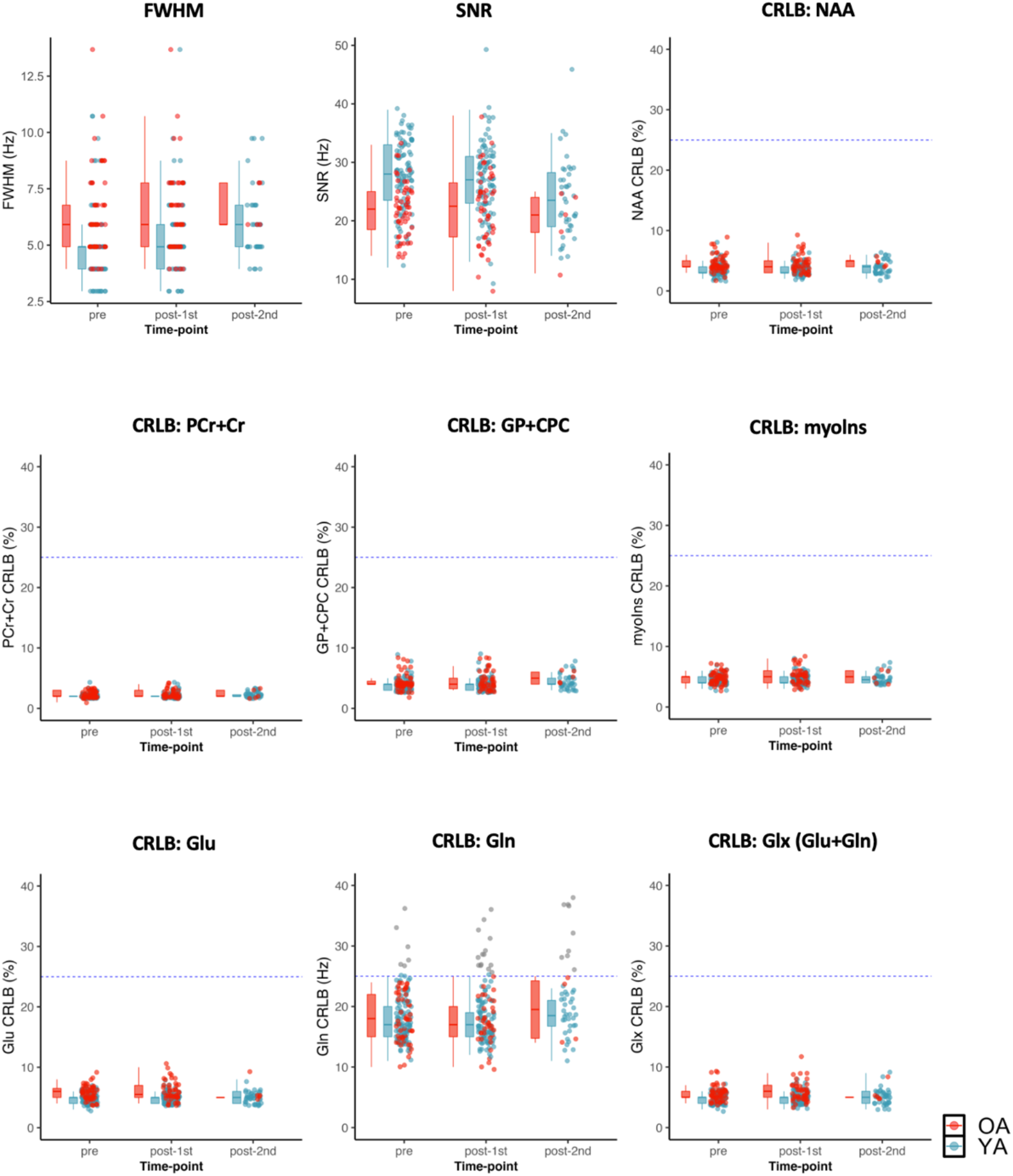
Quality control metrics related to the MRS scans at each time-point. 9 out of 353 spectra were rejected as bad quality and not included in the figure. CRLB >25% are detected as outliers specified by gray color. FWHM: full width at half maximum of singlet peaks; SNR: signal to noise ratio; CRLB: Cramér-Rao lower bound; NAA:N-acetyl-aspartate; PCr+Cr: phosphocreatine plus creatine ; GP+CPC: glycerophosphocholine plus phosphocholine; myoIns: myo-inositol; Glu: glutamate; Gln: glutamine; Glu_Gln: Glu + Gln.

### Resting heart rate data quality assessment

The resting heart rate data was measured during weekly HRV calibration sessions. The pulse was measured with an infrared pulse plethysmograph (ppg) ear sensor and the interbeat interval (IBI) data was extracted after eliminating ectopic beats or other sources of artifacts through a built-in process in emWave pro software. Fig. 9 depicts distributions of the artifact correction rate, mean heart rate, and RMSSD.

**Fig. 9.**
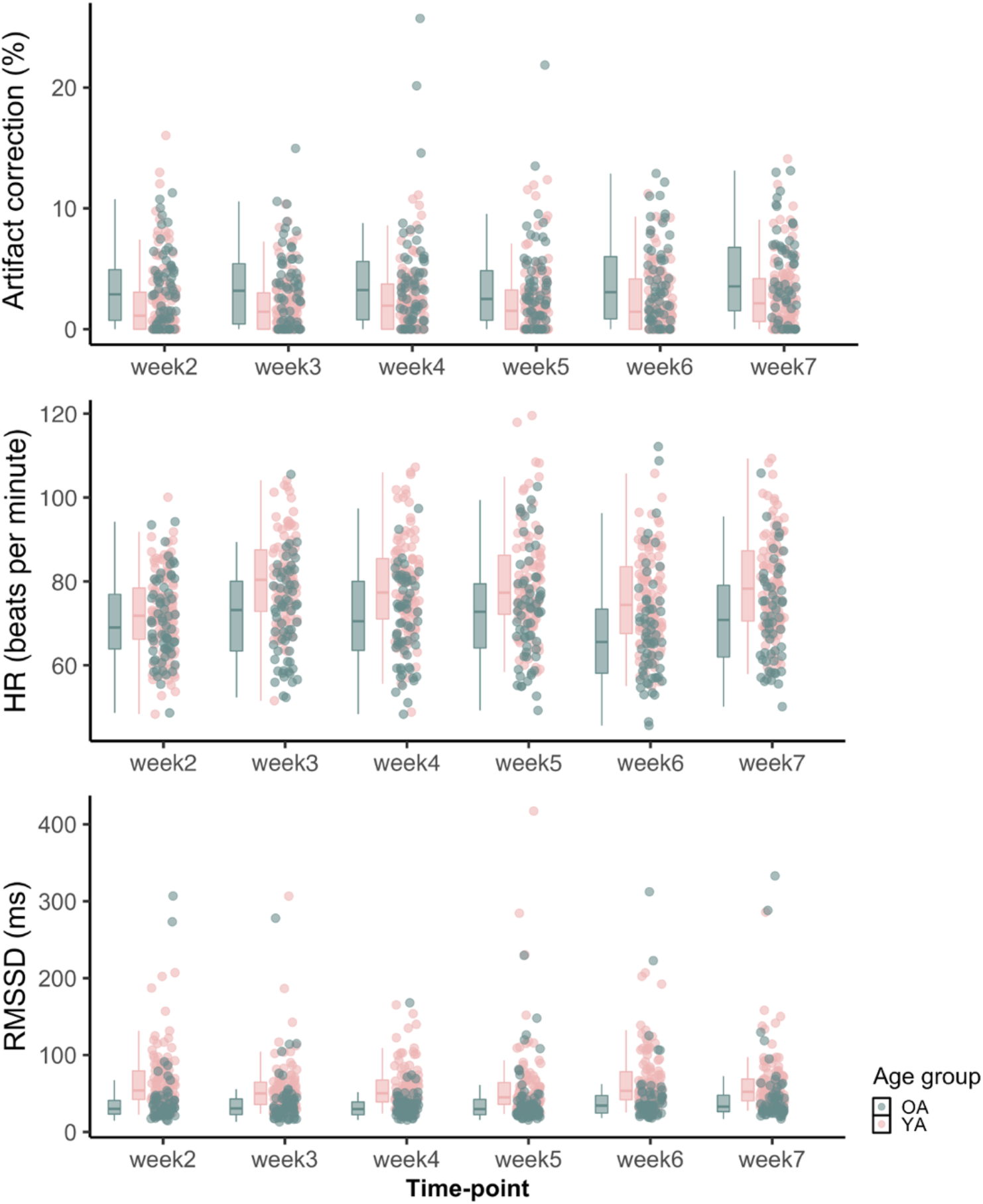
Distribution of artifact correction (%), heart rate (HR), and RMSSD during rest from weekly lab calibration session.

### Behavioral measure quality assessment

We calculated descriptive statistics and reliability estimates of each subscale of the emotion questionnaire (Table 3). We reported average Cronbach’s alpha coefficients from multiple time-points to ensure their internal consistency. To check the test-retest reliability, we reported the intraclass correlation coefficients and typical percentage error^55^.

**Table 3.** Summary of internal consistency (Cronbach’s alpha) and test-retest reliability estimates in emotion questionnaire.

|  | Abbr. | Full Name | Reference | No. of Items | Construct | M (SD) | Cronbach's alpha | Intraclass correlation coefficient | Typical percentage error (%) |
| --- | --- | --- | --- | --- | --- | --- | --- | --- | --- |
| 1 | POMS | The profile of mood states | Grove & Prapavessis (1992) | 40 | Total Mood Disturbance | 88.46 (14.4) | 0.88 (0.85 - 0.91) | 0.67 | 11.02 |
| 2 | SAI | The state anxiety inventory | Spielberger et al.(1983) | 20 | State anxiety | 36.33 (9.36) | 0.72 (0.59 - 0.77) | 0.70 | 15.92 |
| 3 | TAI | The trait anxiety inventory | Spielberger et al.(1983) | 20 | Trait anxiety | 38.46 (10.04) | 0.55 (0.46 - 0.61) | 0.71 | 14.86 |
| 4 | CESD | The Center for Epidemiological Studies Depression Scale | Radloff (1977) | 20 | Depression | 12.44 (7.74) | 0.71 (0.67 - 0.74) | 0.71 | 37.33 |
| 5 | FFMQ | The Five Facet Mindfulness Questionnaire | Tran et al. (2013) | 20 | Mindfulness | 67.98 (10.33) | 0.65 (0.60 - 0.70) | 0.82 | 6.64 |
| 6 | SRSI3 | The Smith Relaxation States Inventory 3-I | Smith (2001) | 38 (26) | Relaxation (state) | 3.11 (0.84) | 0.94 (0.93 - 0.95) | 0.71 | 15.41 |
|  |  |  |  | 38 (8) | Stress (state) | 1.81 (0.64) | 0.83 (0.82- 0.83) | 0.56 | 26.16 |
|  |  | The Smith Relaxation States Inventory 3-II |  | 38 (26) | Relaxation (frequency) | 3.78 (0.85) | 0.93 (0.927- 0.932) | 0.66 | 13.83 |
|  |  |  |  | 38 (8) | Stress (frequency) | 3.1 (0.96) | 0.82 (0.818- 0.826) | 0.72 | 17.33 |
| 7 | CFQ 11 | The Chalder Fatigue Scale | Jackson (2015) | 11 | Fatigue | 12.48 (4.46) | 0.86 (0.85 - 0.87) | 0.56 | 25.53 |
| 8 | ERQ | The Emotion Regulation Questionnaire | Gross & John (2003) | 10 | Emotion regulation (frequency) | 45.40 (7.24) | 0.75 (0.74 - 0.76) | 0.67 | 10.15 |
|  |  |  |  | 10 | Emotion regulation (self-efficacy) | 49.92 (8.96) | 0.88 (0.86 - 0.90) | 0.55 | 13.52 |
| 9 | PSS-10 | Perceived stress scale | Cohen et al., (1983) | 10 | Perceived stress | 2.65 (0.59) | 0.86 (0.84 - 0.89) | 0.61 | 14.61 |

### Physiological measure quality assessment

Table 4 summarizes the main characteristics of physiological data collected during 7-minute resting-state BOLD MRI scans over two time-points. The original data files during MRI scans were stored using the Biopac AcqKnowledge software and later converted as a tsv file for sharing.

**Table 4.** Descriptive statistics and test-retest correlation of physiological measures during resting scan.

| Age group | Condition |  | Respiration rate (Hz) |  | Heart rate (bpm) |  | ETCO<sub>2</sub>(mmHg) |  |
| --- | --- | --- | --- | --- | --- | --- | --- | --- |
|  |  |  | week 2 | week 7 | week 2 | week 7 | week 2 | week 7 |
| YA | Osc+ | M | 0.27 | 0.25 | 68.90 | 66.14 | 41.00 | 40.44 |
|  |  | SD | 0.06 | 0.07 | 9.65 | 8.69 | 3.13 | 3.27 |
|  |  | N | 54 | 46 | 59 | 45 | 35 | 41 |
|  | Osc- | M | 0.29 | 0.29 | 69.79 | 67.97 | 40.56 | 40.76 |
|  |  | SD | 0.06 | 0.06 | 10.33 | 9.65 | 4.02 | 3.99 |
|  |  | N | 43 | 41 | 51 | 43 | 31 | 37 |
| | Week 2 & 7 correlation | | $r(68) = 0.69, p < 0.001$ | | $r(80) = 0.53, p < 0.001$ | | $r(44) = 0.50, p < 0.001$ | |
| OA | Osc+ | M | 0.23 | 0.22 | 65.87 | 62.11 | 39.97 | 40.11 |
|  |  | SD | 0.07 | 0.06 | 9.55 | 10.95 | 4.78 | 4.42 |
|  |  | N | 28 | 25 | 29 | 25 | 28 | 24 |
|  | Osc- | M | 0.22 | 0.23 | 70.35 | 69.34 | 41.07 | 41.65 |
|  |  | SD | 0.06 | 0.06 | 11.78 | 13.59 | 4.04 | 4.30 |
|  |  | N | 24 | 24 | 27 | 21 | 25 | 25 |
| | Week 2 & 7 correlation | | $r(40) = 0.81, p < 0.001$ | | $r(41) = 0.89, p < 0.001$ | | $r(42) = 0.80, p < 0.001$ | |

## Supporting information

CONSORT checklist

## Data Availability

The data described in this paper are available on the OpenNeuro data sharing platform (https://openneuro.org/datasets/ds003823)

## Code Availability

All code for collecting, formatting, and processing the data is available at https://github.com/EmotionCognitionLab/HRV-ER-dataset_release. Information about the code dependencies and package requirements are available in the same Github repository.

## Acknowledgements

This study was supported by NIH R01AG057184 (PI Mather). We thank study participants for their contribution. We thank the research assistant team for their help with data collection: Michelle Wong, Kathryn Cassutt, Collin Amano, Heekyung Rachael Kim, Seungyeon Lee, Alexandra Haydinger, Lauren Thompson, Gabriel Shih, Divya Suri, Sophia Ling, Akanksha Jain, Linette Bagtas, Sumedha Attanti, Ivy Hsu, Michael Kwan, and Juliana Lee.

## Author contributions

The authors made the following contributions. HY: Conceptualization, Data curation, Formal analysis, Investigation, Methodology, Software, Validation, Visualization, Writing - original draft; KN: Conceptualization, Data curation, Investigation, Writing - review & editing, Project administration; JM: Conceptualization, Data curation, Investigation; CCho: Conceptualization, Data curation, Investigation, Resources, Project administration; NM: Data curation, Software, Validation; SB, PN, & SP: Conceptualization, Data curation, Investigation, Writing - review & editing; SD: Investigation, Writing - review & editing; PC, YZ, & VG: Data curation, Investigation; TF: Data curation, Software; JFT, PL, CChang, VM, SN, & DN: Conceptualization, Writing - review & editing; JS: Data curation, Formal analysis, & Writing; JW: Conceptualization & Data curation; MM: Conceptualization, Funding acquisition, Resources, Project administration, Supervision, Writing - review & editing.

